# Evaluation of the diagnostic value of YiDiXie^™^-SS, YiDiXie^™^-HS, and YiDiXie^™^-D in Ovarian cancer

**DOI:** 10.1101/2024.09.15.24313714

**Authors:** Pengwu Zhang, Chen Sun, Zhenjian Ge, Wenkang Chen, Yingqi Li, Shengjie Lin, Wuping Wang, Siwei Chen, Yutong Wu, Huimei Zhou, Xutai Li, Wei Li, Yongqing Lai

## Abstract

**Background:** Ovarian cancer is a serious risk to human health and causes a heavy economic burden. Ultrasound is widely used in the diagnosis of ovarian tumors. However, false-positive ultrasound results bring false diagnosis and wrong surgery or treatment, while false-negative ultrasound results bring missed diagnosis and delayed treatment. There is an urgent need to find convenient, cost-effective and non-invasive diagnostic methods to reduce the false-negative and false-positive rates of ovarian ultrasound. The purpose of this study was to evaluate the diagnostic value of YiDiXie™-SS, YiDiXie™-HS and YiDiXie™-D in Ovarian cancer.

**Patients and methods:** The study finally included 79 study subjects (malignant group, n=12; benign group, n=67). Remaining serum samples from the subjects were collected and tested by applying YiDiXie™ all-cancer detection kit to evaluate the sensitivity and specificity of YiDiXie™-SS, YiDiXie™-HS and YiDiXie™-D, respectively.

**Results:** The sensitivity of YiDiXie™-SS was 100% (95% CI: 75.8% - 100%) and its specificity was 61.2% (95% CI: 49.2% - 72.0%). This means that YiDiXie™ SS has very high sensitivity and high specificity in ovarian tumors. The sensitivity of YiDiXie™ -HS was 83.3% (95% CI: 55.2% - 97.0%) and its specificity was 86.6% (95% CI: 76.4% - 92.8%). This means that YiDiXie™-HS has high sensitivity and high specificity in ovarian tumors.The sensitivity of YiDiXie™-D was 66.7% (95% CI: 39.1% - 86.2%) and its specificity was 92.5% (95% CI: 83.7% - 96.8%). This means that YiDiXie™-D has high sensitivity and very high specificity in ovarian tumors. the sensitivity of YiDiXie™ SS in ultrasound-positive patients was 100% (95% CI: 67.6% - 100%) and its specificity was 61.9% (95% CI: 40.9% - 79.2%). This means that the application of YiDiXie ™ SS reduces the rate of false-positive ovarian ultrasound by 61.9% (95% CI: 40.9% - 79.2%) with essentially no increase in malignant tumor underdiagnosis.The sensitivity of YiDiXie™-HS in ultrasound-negative patients was 75.0% (95% CI: 30.1% - 98.7%) and its specificity was 84.8% (95% CI: 71.8% - 92.4%). This means that the application of YiDiXie™-HS reduces the false negative rate of ultrasound by 75.0% (95% CI: 30.1% - 98.7%).YiDiXie™-D had a sensitivity of 62.5% (95% CI: 30.6% - 86.3%) and a specificity of 90.5% (95% CI: 71.1% - 98.3%) in ultrasound positive patients. This means that YiDiXie™-D reduces the false positive rate of ultrasound by 90.5% (95% CI: 71.1% - 98.3%).The sensitivity of YiDiXie™-D in ultrasound-negative patients was 75.0% (95% CI: 30.1% - 98.7%) and its specificity was 93.5% (95% CI: 82.5% - 97.8%). This means that YiDiXie™-D reduces the false-negative rate of ultrasound by 75.0% (95% CI: 30.1% - 98.7%) while maintaining high specificity.

**Conclusion:** YiDiXie™-SS provides extremely high sensitivity and relatively high specificity in ovarian tumors.YiDiXie™-HS provides high sensitivity and high specificity in ovarian tumors.YiDiXie™-D provides high sensitivity and extremely high specificity in ovarian tumors.YiDiXie™-SS significantly reduces false-positive rates on ovarian ultrasound with essentially no increase in delayed treatment of malignancies. YiDiXie™-HS significantly reduces the false-negative rate of ovarian ultrasound, and YiDiXie™-D can significantly reduce the false-positive rate of ovarian ultrasound or significantly reduce the false-negative rate of ovarian ultrasound while maintaining a high level of specificity. The YiDiXie ™ test has significant diagnostic value in ovarian cancer, and is expected to solve the two problems of the “too high false-positive rate” and the “too high false-negative rate” of ovarian ultrasound.

**Clinical trial number:** ChiCTR2200066840.

## INTRODUCTION

Ovarian cancer is one of the common malignancies in gynecology[1, 2]. The latest data shows that there will be 324,398 new cases of ovarian malignant tumors and 206,839 new deaths worldwide in 2022[3]. Compared with 2020, the incidence of ovarian malignant tumors increased by 3.32% in 2022, still showing an increasing trend every year[4]. Studies have shown that approximately 80% of Ovarian cancer are diagnosed in the intermediate or late stages due to the lack of specific signs and symptoms and the lack of reliable screening for early Ovarian cancer detection[5, 6]. Unfortunately, the 5-year survival rate for patients with stage III and IV Ovarian cancer is only 20%[7]. However, the 5-year survival rate for patients with stage I and II Ovarian cancer rises to 70-90%[8,9]. Studies have shown that Ovarian cancer have the highest mortality rate of all gynecologic malignancies[10]. Early detection not only improves patient prognosis, but also reduces the financial burden on patients[11,12]. Therefore, ovarian malignant tumors are a serious threat to human health[13].

Ultrasound is widely used in the diagnosis of ovarian tumors[14]. On the one hand, ultrasound can produce a large number of false positive results. Studies have shown that ultrasound has a false positive rate of 23.3% in the diagnosis of Ovarian cancer[15]. Patients usually undergo radical surgery when the ultrasound is positive[16, 17]. False-positive ultrasound results mean that a benign disease is misdiagnosed as a malignant tumor. The patient will probably have to bear the adverse consequences of unnecessary mental suffering, expensive surgery and investigations, surgical trauma, organ removal, loss of function, and even serious perioperative complications[18]. Therefore, there is an urgent necessity to find a convenient, cost-effective and noninvasive diagnostic method to reduce the false positive rate of ovarian ultrasound.

On the other hand, ultrasound can produce a great number of false-negative results. Studies have shown that ultrasound has a false negative rate of 42.9% in the diagnosis of Ovarian cancer[19]. Patients are usually taken for observation and regular follow-up when the ultrasound is negative[16, 17]. False-negative ultrasound results imply misdiagnosis of a malignant tumor as a benign condition, which will likely lead to delayed treatment, progression of the malignant tumor, and possibly even development of an advanced stage[20]. Patients will have to bear the adverse consequences of poor prognosis, expensive treatment, poor quality of life and short survival as a result[21]. Thus, there is an urgent need to find a convenient, cost-effective and noninvasive diagnostic method to reduce the false-negative rate of ovarian ultrasound.

In addition, there are some special patients who need to be extra cautious in choosing whether or not to operate, such as: smaller tumors, strong fertility requirements, ovarian insufficiency, and poor general condition of the patient. The risk of wrong surgery in these special patients is much higher than the risk of missed diagnosis. And false-positive ultrasound results mean that benign diseases are misdiagnosed as malignant tumors, which will lead to misdiagnosis and wrong surgery. Therefore, there is an urgent need to find a convenient, cost-effective and noninvasive diagnostic method with high specificity to substantially reduce the false-positive rate of ovarian ultrasound in these special patients or to significantly reduce the false-negative rate while maintaining high specificity.

Based on the detection of novel tumor markers of miRNA in serum, an in vitro-diagnostic test product, the YiDiXie ™ all-cancer test (hereinafter referred to as the YiDiXie™ test, the YiDiXie™ test), has been developed by Shenzhen KeRuiDa Health Technology Company Limited[22]. The YiDiXie™ test is an in vitro diagnostic product that detects a wide range of cancer types in as little as 200 microliters of whole blood or 100 microliters of serum per test[22]. The YiDiXie™ test consists of three different products with different performance profiles: the YiDiXie™-HS, YiDiXie™-SS, and the YiDiXie™-D[22].

The aim of this study was to estimate the diagnostic value of YiDiXie ™ -SS, YiDiXie ™ -HS and YiDiXie™-D in Ovarian cancer.

## PATIENTS AND METHODS

### Study design

This work is part of the sub-study “Evaluating the value of the YiDiXie ™ test as an adjuvant diagnostic in multiple tumors” of the SZ-PILOT study (ChiMRIR2200066840).

The SZ-PILOT study (ChiMRIR2200066840) is a single-center, prospective, observational study. Subjects who signed a pan informed consent for donation of remaining samples at the time of admission or physical examination were included, and 0.5 ml of their remained serum samples were collected for use in this study.

The study was blinded. Neither the laboratory personnel who performed the YiDiXie ™ test nor the KeRuiDa laboratory technicians who determined the results of the YiDiXie™ test were aware of the subjects’ clinical information. The clinical experts who evaluated the subjects’ clinical information were also unaware of the results of the YiDiXie™ test.

The research was approved by the Ethics Committee of Shenzhen Hospital of Peking University and was conducted in accordance with the International Conference on Harmonization (ICH) Code of Practice for the Quality Management of Pharmaceutical Clinical Trials and the Declaration of Helsinki.

### Participants

The two groups of participants were enrolled separately, and all subjects who satisfied the inclusion criteria were included consecutively.

This research initially included hospitalized patients with “suspected (solid or hematological) malignancy” who signed a general informed consent for donation of the remaining samples. Subjects with a postoperative pathologic diagnosis of “malignant tumor” were included in the malignant group, and those with a postoperative pathologic diagnosis of “benign disease” were included in the benign group. Subjects with unclear pathology results were excluded from the study. A portion of the samples from the malignant group were used in our previous article [22].

Subjects who failed the serum sample quality test prior to the YiDiXie™ test were excluded from this study. Refer to our previous article for specific enrollment and exclusion[22].

### Sample collection, processing

The serum samples used in this study were obtained from serum left over after a normal clinic visit, without the need for additional blood draws. Approximately 0.5 ml of serum samples were collected from the remaining serum of subjects in the Medical Laboratory Department and stored at -80°C for use in the subsequent the YiDiXie™ test.

### The YiDiXie test

The YiDiXie ™ test was performed using the YiDiXie ™ all-cancer detection kit, an in vitro diagnostic kit developed and manufactured by Shenzhen KeRuiDa Health Technology Co. for use in fluorescent quantitative PCR instruments. It detects the expression level of dozens of miRNA biomarkers in the serum to determine whether cancer exists in the body of the test subject[22]. It predefines appropriate thresholds for each miRNA biomarker to ensure that each miRNA marker is highly specific, and integrates these independent assays in a concurrent testing format to significantly increase the sensitivity and maintain the specificity of a broad spectrum of cancers[22].

The YiDiXie™ test consists of three assays with very different characteristics: YiDiXie™-HS, YiDiXie ™-SS, and YiDiXie™-D[22]. YiDiXie™-HS (YiDiXie™ -Highly Sensitive, YiDiXie ™ -HS) has been developed with a balance of sensitivity and specificity[22]. YiDiXie ™ -SS (YiDiXie ™ -Super Sensitive, YiDiXie ™ -SS) has significantly increased the number of miRNA assays to achieve extremely high sensitivity for all clinical stages of all malignant tumor types[22]. YiDiXie™-D (YiDiXie™-Diagnosis, YiDiXie ™ -D) dramatically increases the diagnostic threshold of individual miRNA tests to achieve extremely high specificity for the full range of malignant tumor types[22].

Perform the YiDiXie ™ test according to the instruction manual of the YiDiXie ™ all-cancer detection kit. Refer to our previous article for detailed procedures[22].

The original test results were analyzed by the laboratory technicians of Shenzhen KeRuiDa Health Technology Company Limited and the results of the YiDiXie™ test were determined to be “positive” or “negative”[22].

### Ultrasound diagnosis

The diagnostic conclusion of the ultrasound is used to determine whether the result is “positive” or “negative”. If the diagnostic conclusion is positive, more positive, or favors malignant tumors, the test result is determined to be “positive”. A diagnosis that is positive, more positive or favors a benign disease, or a diagnosis that is ambiguous as to whether it is benign or malignant, is considered “negative”.

### Extraction of clinical data

Clinical, pathological, laboratory, and imaging data in this study were obtained from the participants’ hospitalized medical records or physical examination reports. Clinical staging was completed by trained clinicians assessed according to the AJCC staging manual (7th or 8th edition)[23, 24].

### Statistical analyses

For demographic and baseline characteristics, descriptive statistics were reported. For categorical variables, the number and percentage of subjects in each category were calculated. For continuous variables, the total number of subjects (n), mean, standard deviation (SD) or standard error (SE), median, first quartile (Q1), third quartile (Q3), minimum, and maximum values were calculated. 95% confidence intervals (CI) for multiple indicators were calculated using the Wilson (score) method.

## RESULTS

### Participant disposition

The study finally included 79 study subjects (malignant group, n=12; benign group, n=67). The demographic and clinical characteristics of the 79 study subjects are presented in Table 1.

**Table 1.**
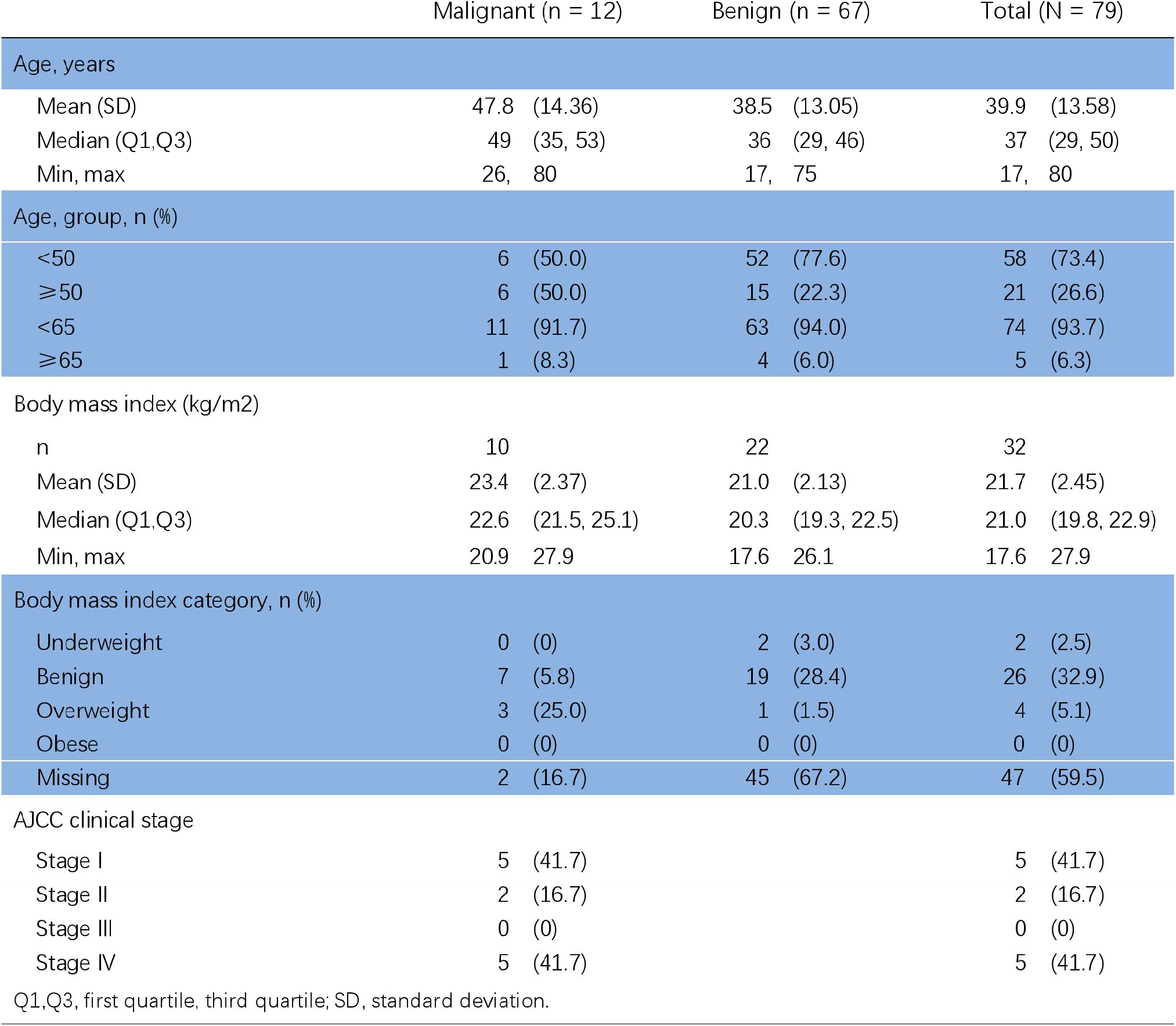
Participants’ demographic and clinical manifestation.

The two groups of study subjects were comparable in terms of demographic and clinical characteristics (Table 1). The mean (standard deviation) age was 39.9 (13.58) years.

### Diagnostic performance of YiDiXie™-SS

As shown in Table 2, the sensitivity of YiDiXie™ -SS was 100% (95% CI: 75.8% - 100%) and its specificity was 61.2% (95% CI: 49.2% - 72.0%). This means that YiDiXie ™ SS has very high sensitivity and high specificity in ovarian tumors.

**Table 2.** Performance of YiDiXie™-SS.

|  | Malignant | Benign | Total |
| --- | --- | --- | --- |
|  | 12 | 67 | 79 |
| Positive | 12 | 26 | 36 |
| Negative | 0 | 41 | 41 |
| SEN = 12/12 = 100% (75.8% - 100%) SPE = 41/67 = 61.2% (49.2% - 72.0%) |  |  |  |

Two-sided 95% Wilson confidence intervals were calculated. SEN, Sensitivity. SPE, Specificity.

### Diagnostic performance of YiDiXie™-HS

As shown in Table 3, the sensitivity of YiDiXie™ -HS was 83.3% (95% CI: 55.2% - 97.0%) and its specificity was 86.6% (95% CI: 76.4% - 92.8%). This means that YiDiXie ™ -HS has high sensitivity and high specificity in ovarian tumors.

**Table 3.** Performance of YiDiXie™-HS.

|  | Malignant | Benign | Total |
| --- | --- | --- | --- |
|  | 12 | 67 | 79 |
| Positive | 10 | 9 | 19 |
| Negative | 2 | 58 | 60 |
| SEN = 10/12 = 83.3% (55.2% - 97.0%) SPE = 58/67 = 86.6% (76.4% - 92.8%) |  |  |  |

Two-sided 95% Wilson confidence intervals were calculated. SEN, Sensitivity. SPE, Specificity.

### Diagnostic performance of YiDiXie™-D

As shown in Table 4, the sensitivity of YiDiXie™ -D was 66.7% (95% CI: 39.1% - 86.2%) and its specificity was 92.5% (95% CI: 83.7% - 96.8%). This means that YiDiXie ™ -D has high sensitivity and very high specificity in ovarian tumors.

**Table 4.** Performance of YiDiXie™-D.

|  | Malignant | Benign | Total |
| --- | --- | --- | --- |
|  | 12 | 67 | 79 |
| Positive | 8 | 5 | 13 |
| Negative | 4 | 62 | 66 |
| SEN = 8/12 = 66.7% (39.1% - 86.2%) SPE = 62/67 = 92.5% (83.7% - 96.8%) |  |  |  |

Two-sided 95% Wilson confidence intervals were calculated. SEN, Sensitivity. SPE, Specificity.

### Diagnostic performance of ultrasound

As indicated in Table 5, the ultrasound sensitivity was 66.7% (95% CI: 39.1% - 86.2%) and its specificity was 68.7% (95% CI: 56.8% - 78.5%).

**Table 5.**
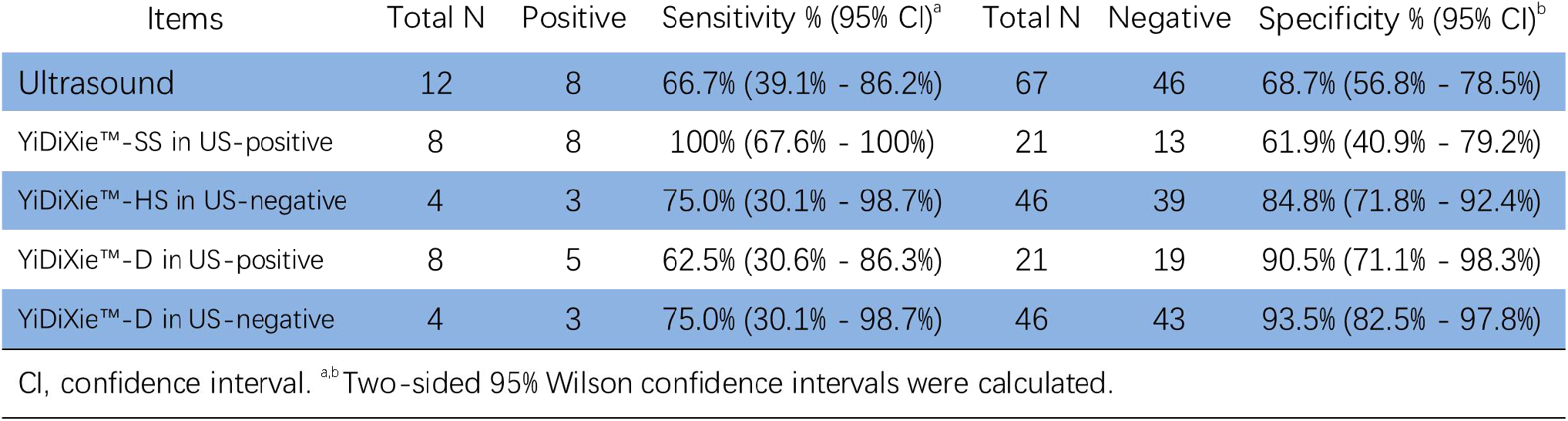
Performance of different Items.

### Diagnostic performance of YiDiXie™-SS in ovary ultrasound-positive patients

To address the challenge of high false positive rate of ovarian ultrasound, YiDiXie ™ -SS was applied to ovarian ultrasound positive patients.

As shown in Table 5, the sensitivity of YiDiXie™ SS in ultrasound-positive patients was 100% (95% CI: 67.6% - 100%) and its specificity was 61.9% (95% CI: 40.9% - 79.2%). This means that the application of YiDiXie ™ SS reduces the rate of false-positive ovarian ultrasound by 61.9% (95% CI: 40.9% - 79.2%) with essentially no increase in malignant tumor underdiagnosis.

### The Diagnostic performance of YiDiXie™-HS in ovary ultrasound-negative patients

To resolve the challenge of high ovarian ultrasound leakage rate, YiDiXie™-HS was applied to ultrasound-negative patients.

As shown in Table 5, the sensitivity of YiDiXie™ -HS in ultrasound-negative patients was 75.0% (95% CI: 30.1% - 98.7%) and its specificity was 84.8% (95% CI: 71.8% - 92.4%). This means that the application of YiDiXie™-HS reduces the false negative rate of ultrasound by 75.0% (95% CI: 30.1% - 98.7%).

### The Diagnostic performance of YiDiXie™-D in ovary ultrasound-positive patients

The consequences of false-positive ultrasound were significantly worse than those of false-negative ultrasound in some ultrasound-positive patients, so YiDiXie ™ -D was applied to such patients to reduce their false-positive rate.

As shown in Table 5, YiDiXie ™ -D had a sensitivity of 62.5% (95% CI: 30.6% - 86.3%) and a specificity of 90.5% (95% CI: 71.1% - 98.3%) in ultrasound positive patients. This means that YiDiXie TM -D reduces the false positive rate of ultrasound by 90.5% (95% CI: 71.1% - 98.3%).

### The Diagnostic performance of YiDiXie™-D in ovary ultrasound-negative patients

Certain ultrasound-negative patients had significantly more severe false-positive than false-negative consequences, so the more specific YiDiXie™-D was applied to such patients.

As shown in Table 5, the sensitivity of YiDiXie™ -D in ultrasound-negative patients was 75.0% (95% CI: 30.1% - 98.7%) and its specificity was 93.5% (95% CI: 82.5% - 97.8%). This means that YiDiXie ™ -D reduces the false-negative rate of ultrasound by 75.0% (95% CI: 30.1% - 98.7%) while maintaining high specificity.

## DISCUSSION

### Clinical significance of YiDiXie™-SS in ovary ultrasound-positive patients

In patients with positive ovarian ultrasound, further diagnostic methods are both important in terms of sensitivity and specificity. Higher false-negative rates mean that more Ovarian cancer are underdiagnosed, which will lead to delays in their treatment, progression of the malignancy, and possibly even development of advanced stages. A higher false-positive rate means that more benign ovarian diseases are misdiagnosed and will likely lead to unnecessary surgery. In general, misdiagnosis of benign ovarian diseases as malignant tumors will usually lead to unnecessary surgery, but it will not affect the patient’s prognosis, and its treatment cost is much lower than that of advanced cancer.

In addition, the positive predictive value is higher in patients with positive ovarian ultrasound. This makes the false-negative rate more harmful even if it is comparable to the false-positive rate. Therefore, YiDiXie™-SS, an extremely high sensitivity but slightly lower specificity, was chosen for reducing the false positive rate of ovarian ultrasound.

As indicated in Table 5, YiDiXie ™ -SS had a sensitivity of 100% (95% CI: 67.6% - 100%) and its specificity was 61.9% (95% CI: 40.9% - 79.2%) in ultrasound-positive patients. The above results indicate that YiDiXie™-SS reduces the false positive rate of ovarian ultrasound by 61.9% (95% CI: 40.9% - 79.2%) while maintaining a sensitivity close to 100%.

This means that YiDiXie ™ -SS substantially reduces the probability of misdiagnosis of benign ovarian disease as malignancy with essentially no increase in delayed treatment of malignancy. Thus, YiDiXie™-SS meets the clinical needs well and has significant clinical implications in Ovarian cancer.

### Clinical implications of YiDiXie™-HS in ovary ultrasound-negative patients

In patients with negative ovarian ultrasound, further diagnostic approaches are important in terms of both sensitivity and specificity. Higher false-negative rates mean that more Ovarian cancer are underdiagnosed, which will lead to delays in their treatment, progression of the malignancy, and possibly even development of advanced stages. In general, when benign ovarian disease is misdiagnosed as a malignant tumor, the patient usually undergoes surgery, which does not affect the patient’s prognosis, and the cost of its treatment is much lower than that of advanced cancer. Therefore, in patients with negative ovarian ultrasound, the higher false-negative rate can lead to significant harm.

In addition, the negative predictive value is higher in patients with negative ovarian ultrasound. Thus higher false positive rates can lead to significant harm. Therefore, YiDiXie™-HS with high sensitivity and specificity was selected for reducing the false negative rate of ovarian ultrasound.

As indicated in Table 5, the sensitivity of YiDiXie ™ -HS in ultrasound-negative patients was 75.0% (95% CI: 30.1% - 98.7%) and its specificity was 84.8% (95% CI:71.8% - 92.4%). The above results indicate that the application of YiDiXie ™ -HS reduced the false-negative rate of ultrasound by 75.0% (95% CI: 30.1% - 98.7%).

This means that YiDiXie ™ -HS substantially reduces the probability of negative ovarian ultrasound for missed malignancies. Therefore, YiDiXie™-HS meets the clinical needs well and has important clinical implications in Ovarian cancer.

### Clinical significance of YiDiXie™-D

Ovarian tumors considered malignant usually undergo radical surgery. However, some patients require extra caution in choosing whether or not to undergo surgery and therefore require further diagnosis, e.g., smaller tumors, strong fertility requirements, ovarian insufficiency, poor general condition of the patient, etc.

In patients with ovarian tumors, both the sensitivity and specificity of further diagnostic methods are important. The trade-off between sensitivity and specificity is essentially a trade-off between the “risk of underdiagnosis of Ovarian cancer” and the “risk of misdiagnosis of benign ovarian disease”. Because smaller tumors have a lower risk of tumor progression and distant metastasis, the “risk of missed diagnosis of Ovarian cancer” is much lower than the “risk of misdiagnosis of benign ovarian disease”. For patients with poor general conditions, the “risk of misdiagnosis of benign ovarian disease” is much higher than the “risk of misdiagnosis of malignant ovarian tumor” because the perioperative risk is much higher than the general condition. Therefore, for these patients, YiDiXie™-D, which has very high specificity but low sensitivity, was chosen to reduce the rate of false-positive ovarian ultrasound or to significantly reduce the rate of false-negative ovarian ultrasound while maintaining high specificity.

As shown in Table 5, the sensitivity of YiDiXie™ -D in ultrasound-positive patients was 62.5% (95% CI: 30.6% - 86.3%), and its specificity was 90.5% (95% CI: 71.1% - 98.3%), while the sensitivity of YiDiXie™ -D in ultrasound-negative patients was 75.0% (95% CI: 30.1% - 98.7%), and its specificity was 93.5% (95% CI: 82.5% - 97.8%). This means that YiDiXie ™ -D reduces the false-positive rate of ultrasound by 90.5% (95% CI: 71.1% - 98.3%), or reduces the false-negative rate of ultrasound by 75.0% (95% CI: 30.1% - 98.7%) while maintaining a high specificity.

This implies that YiDiXie ™ -D substantially reduces the probability of wrong surgery in these patients with ovarian tumors that require extra cautious surgery. Thus, YiDiXie ™ -D meets the clinical needs well and has important clinical implications in Ovarian cancer.

### The YiDiXie™ test is promising to address two challenges of ovarian tumors

Firstly, YiDiXie ™ -SS significantly reduces the risk of misdiagnosis of benign ovarian disease as Ovarian cancer. On the one hand, YiDiXie ™ -SS substantially reduces the probability of incorrect surgery for benign ovarian diseases with essentially no increase in missed diagnosis of ovarian malignant tumors. As illustrated in Table 5, YiDiXie™ -SS reduced the rate of false-positive ovarian ultrasound by 61.9% (95% CI: 40.9% - 79.2%) with essentially no increase in missed diagnosis of Ovarian cancer. Hence, YiDiXie ™ -SS significantly reduces a number of adverse outcomes associated with unnecessary surgery with essentially no increase in delayed treatment of Ovarian cancer.

On the other hand, YiDiXie ™ -SS greatly relieves surgeons from non-essential work. The patient usually undergoes radical surgery when the ovaries are ultrasound positive. The timely completion of these radical surgeries is directly dependent on the number of surgeons. In many parts of the world, appointments are made for months or even more than a year. This inevitably delays the treatment of Ovarian cancer therein, and thus it is not uncommon for ultrasound-positive ovarian patients awaiting radical surgery to experience malignancy progression or even distant metastases. As illustrated in Table 6, YiDiXie ™ -SS reduced the rate of false-positive ovarian ultrasound by 61.9% (95% CI: 40.9% - 79.2%) with essentially no increase in the number of missed diagnoses of ovarian malignant tumors. As a conclusion, YiDiXie ™ -SS can greatly relieve surgeons of non-essential workloads and facilitate the timely diagnosis and treatment of Ovarian cancer or other conditions that would otherwise be delayed.

Secondly, YiDiXie™-HS greatly reduces the risk of missed diagnosis of Ovarian cancer. Ovarian malignant tumors are usually temporarily excluded when ovarian ultrasound is negative. The high rate of false-negative ovarian ultrasound thus leads to delayed treatment for a large number of patients with ovarian malignant tumors. As illustrated in Table 7, YiDiXie ™ -HS reduced the false-negative rate of ovarian ultrasound by 75.0% (95% CI: 30.1% - 98.7%). Therefore, YiDiXie ™ -HS substantially reduced the probability of missed malignant tumor diagnosis by ovarian ultrasound and facilitated timely diagnosis and treatment of patients with ovarian malignant tumors who would otherwise have been delayed in treatment.

Thirdly, YiDiXie ™ -D is expected to solve the problem of “ high false positive rate “ and “ high false negative rate “ in some specific patients. Ovarian tumors are usually treated with radical surgery when malignancy is considered. However, in cases such as smaller tumors, insufficient ovarian function and poor general condition of the patient, surgery may result in unnecessary surgical trauma, organ removal, loss of function, or serious perioperative complications. Thus, these patients require extra caution before surgery. As illustrated in Table 5, YiDiXie™ D reduced the false-positive ultrasound rate by 90.5% (95% CI: 71.1% - 98.3%) or, while maintaining high specificity, reduced the false-negative ultrasound rate by 75.0% (95% CI: 30.1% - 98.7%). Thus, YiDiXie ™ -D substantially reduced the risk of wrong surgery or serious perioperative complications in these patients.

Finally, the YiDiXie ™ test allows for “just-in-time” diagnosis of ovarian tumors. On the one hand, the YiDiXie ™ test requires only a tiny amount of blood, allowing patients to complete the diagnostic process non-invasively without having to leave their homes. 0.2 mL of whole blood is sufficient to complete The YiDiXie™ test [22]. The average patient can collect 0.2 mL of finger blood at home using a finger blood collection needle without the need for a medical staff to collect blood from a vein [22]. Therefore, patients can complete the diagnostic process non-invasively without leaving their homes.

On the other hand, the YiDiXie ™ test has a nearly unlimited diagnostic capacity. Figure 1 shows the basic flowchart of the YiDiXie ™ test, from which it can be seen that the YiDiXie™ test does not require either a doctor or medical equipment, nor does it require medical personnel to collect blood. As a result, the YiDiXie ™ test is completely independent of the number of medical personnel and medical facilities, and its testing capacity is nearly unlimited. As a result, the YiDiXie™ test enables “just-in-time” diagnosis of ovarian tumors without patients having to wait anxiously for an appointment.

**Figure 1.**
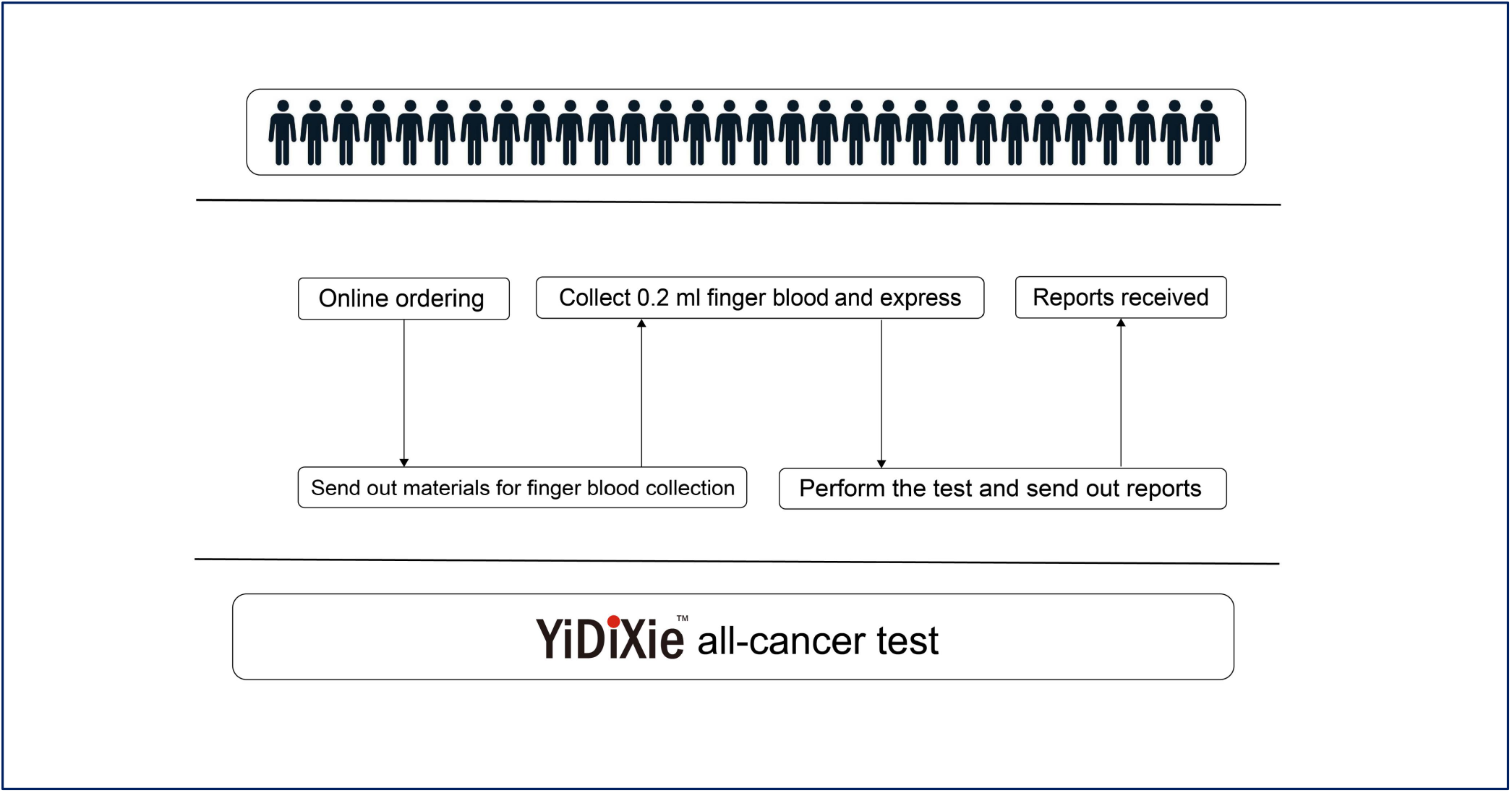
Basic flowchart of the YiDiXie™ test.

In short, the YiDiXie™ test has significant diagnostic value in ovarian cancer, and has the potential to solve the “high false-positive rate” and “high false-negative rate” of ovarian tumors.

### Limitations of the study

Firstly, the number of cases in this research was small and future clinical studies with a larger sample size are needed for further evaluation.

Secondly, this was an inpatient case-benign disease-controlled study of malignant tumors, and future cohort studies in the natural population of ovarian tumors are needed for further evaluation. Finally, this study was a single-center study, which may have led to some degree of bias in the results of this study. The future multicenter study is needed to further evaluate it.

## CONCLUSION

YiDiXie ™ -SS provides extremely high sensitivity and relatively high specificity in ovarian tumors.YiDiXie ™ -HS provides high sensitivity and high specificity in ovarian tumors.YiDiXie ™ -D provides high sensitivity and extremely high specificity in ovarian tumors.YiDiXie ™ -SS significantly reduces false-positive rates on ovarian ultrasound with essentially no increase in delayed treatment of malignancies. YiDiXie ™ -HS significantly reduces the false-negative rate of ovarian ultrasound, and YiDiXie ™ -D can significantly reduce the false-positive rate of ovarian ultrasound or significantly reduce the false-negative rate of ovarian ultrasound while maintaining a high level of specificity. The YiDiXie™ test has significant diagnostic value in ovarian cancer, and is expected to solve the two problems of the “too high false-positive rate” and the “too high false-negative rate” of ovarian ultrasound.

## Data Availability

All data produced in the present study are contained in the manuscript.

